# Integrating Multi-Ancestry Polygenic Risk Scores and Wearable Data to Detect Depression and Gene-Environment Interactions in Youth

**DOI:** 10.1101/2025.11.27.25341118

**Authors:** Nathan Chen, Mahnoor Hyat, Jinhan Zhu, Anind Dey, Jennifer K. Forsyth

## Abstract

**Background:** Depression (DEP) has an estimated heritability of ∼37%. Polygenic risk scores (PRS), which aggregate common genetic variant effects, account for up to 8.4% variance in case versus control status. Understanding the relationships between sleep, activity, and DEP symptoms, and whether these relationships differ depending on genetic predisposition is crucial for early identification, particularly during adolescence when DEP rates tend to increase.

**Method:** 2,768 adolescents (African [AFR], American Admixed [AMR], and European [EUR] ancestry) from the Adolescent Brain Cognitive Development study were examined. DEP-PRS was calculated using PRS-CSx. Average and variance sleep and activity measures were derived from Fitbit wearable data. Mixed effects models examined main and interaction effects of DEP-PRS and each Fitbit measure with DEP severity.

**Results:** DEP-PRS, mean resting heart rate, mean sedentary minutes, and sleep minutes variance were positively associated with DEP severity; mean intense active minutes, total steps, and sleep minutes were negatively associated. DEP-PRS showed interactions with sleep minutes variance, mean total steps, and mean intense active minutes, with stronger associations between these variables in individuals at higher genetic risk. In individuals at higher genetic risk with different ancestries, the association between mean sleep minutes and DEP severity was positive for AFR, negative for AMR, and non-significant for EUR.

**Conclusion:** The relationship between sleep, activity, and DEP severity appears to depend on genetic predisposition Further research with optimized PRS, research-grade wearables, and longitudinal design is needed.

## Introduction

Depression (DEP) is a leading cause of illness and disability among adolescents but remains a frequently underrecognized mental health concern (Thapar 2012). Detecting DEP in youth is often complicated by the predominantly internalizing nature of symptoms, such as anhedonia, fatigue, and difficulty concentrating, which may not be readily apparent to parents, educators, or clinicians (Black 2022). Yet, as untreated depression can result in significant morbidity, heightened risk of self-harm, and impaired school and social functioning during a critical period of development, early detection is critical for facilitating prevention strategies and halting potential negative sequelae.

DEP risk is significantly influenced by both environmental and genetic factors, with twin studies measuring heritability at around 40 percent (Sullivan et al., 2000). Genome-wide association studies (GWAS) have now identified hundreds of common genetic variants that are associated with DEP. However, polygenic risk scores (PRS) for DEP, which estimate an individual’s genetic risk for the disorder as a weighted sum of common risk variants, indicate that PRS can only explain up to 8.7% of the variance in DEP case-control status (Wray et al., 2018). Modest predictive accuracy of DEP PRS likely reflects the partial and strong contributions of environmental factors to DEP. Moreover, given that European ancestry individuals are strongly overrepresented in GWAS of DEP and allele frequencies and linkage disequilibrium patterns vary across global populations, DEP PRS show their highest predictive accuracy in individuals of European ancestry, with lower accuracy in underrepresented populations. This raises critical healthcare equity concerns (Martin et al., 2019). Thus, while PRS holds potential for improving early DEP risk prediction, the current clinical utility of PRS remains limited, emphasizing the need for more inclusive and comprehensive approaches to DEP risk prediction.

One potential complementary path to elucidate the risk factors of DEP alongside PRS is to incorporate measurements of physical activity and sleep (Firth et al., 2020). Decreases in physical activity and increases in sedentary behavior are associated with increased risk of DEP in adolescents (Wang & Pieper, 2022; Hou et al., 2024). Sleep disturbances often precede DEP onset, which can in turn worsen sleep problems in a bidirectional manner (Goldstone et al., 2020, Lovato & Gradisar, 2014; Bacaro et al., 2024). However, a challenge in validating associations between activity and sleep relative to DEP is the discrepancy between self-report and wearable measures of activity and sleep (Kiss et al., 2025; Cerin et al., 2016; Prince et al., 2008). In healthy populations, systematic reviews indicate wearable measures of total sleep time, wake after sleep onset, resting heart rate, and total steps have no significant differences compared to polysomnography (PSG) and research-grade actigraphy devices (Haghayegh et al., 2019; Fuller et al., 2020). Meanwhile, several reviews have examined the lack of agreement between self-reports and PSG measures of total sleep and sleep onset latency, which has also been debated as a construct named sleep misperception (Harvey & Tang, 2012; Castelnovo et al., 2019). Other studies suggest the confounding effects of mood may explain the stronger associations between self-report sleep and DEP symptoms compared to wearable measures (George et al., 2019; Akre et al., 2025). Importantly, multiple studies have found longitudinal associations between wearable measures and DEP and its potential in remote monitoring and patient feedback applications (Nelson et al., 2022; Wainberg et al., 2021; Zhang et al., 2021; Borghare et al., 2024). Wearable measures may also capture a variety of dimensional phenotypes including resting heart rate, step count, sleep variability, and macro sleep architecture, which may further elucidate associations between PRS and granular behavior measures.

In addition to wearable measures of activity and sleep potentially having utility for early detection of DEP, they may also be influenced by genetic risk for DEP, and their utility for early detection of DEP may depend on genetic risk. In one study that examined an adult population without psychiatric conditions, DEP PRS was associated with wearable measures including reduced walking and moderate activities and increased sedentary and sleep time (Dennison et al., 2021). However, the association between PRS and wearable measures for adolescents is unclear. One study found significant positive correlations between DEP PRS and parent-reported measures of disorders of initiating and maintaining sleep and disorders of excessive somnolence for adolescents (Ohi et al., 2021). Another study found that elevated risk for DEP based on having a parent with DEP was associated with average of 26 minutes more total sleep on weeknights than controls; however, familial-based risk of DEP may capture environmental influences to a greater extent than DEP PRS (Wescott et al., 2019). Two recent studies using self-report measures of sleep and activity in adults found that higher levels of recreational activity and sleep durations of 7 to 9 hours grouped with other health lifestyle factors, respectively, were protective of DEP severity regardless of genetic predisposition (Choi et al., 2020; Zhao et al., 2023). However, these studies utilized self-report lifestyle data from one baseline questionnaire and did not examine non-European populations due to the limitations of the selected PRS methods. Furthermore, it is unclear if similar relationships exist in adolescents. Therefore, investigating the associations between DEP PRS, wearable-derived activity and sleep measures, and DEP severity among adolescents with diverse ancestry, is a crucial next step for the field.

Using data from the ABCD Study, a U.S.-based large-scale study on adolescent development with genomic, clinical, demographic and Fitbit wearable data, this study therefore used a multi-ancestry optimized DEP PRS to examine whether: 1) DEP PRS is associated with wearable measures of activity and sleep in individuals without DEP; 2) whether activity and sleep patterns are differentially associated with current DEP severity depending on DEP PRS; 3) and whether these associations are concordant or discordant across different ancestries.

## Methods

### Participants and Ethical Considerations

Data from the ABCD study were used for the present investigation, which is a general population cohort of 11,878 children ages 8–11 years recruited from 21 sites throughout the U.S. (Barch et al., 2018). Each recruitment site obtained assent and consent from the children and their parent(s)/legal guardian(s), respectively, in accordance with local Institutional Reviews Boards. Data analyzed for the present study involved individuals of European (EUR), African (AFR), or American Admixed ancestry (AMR), and with valid Fitbit physical activity and sleep data, resulting in 2,768 unique participants at year 2. Year 2 data was used to obtain Fitbit wearable data from the full ABCD Fitbit protocol. Data was obtained from Release 5.1 from the ABCD consortium.

### Depression History and Symptom Severity

Lifetime history of DEP (Lifetime-DEP) was assessed using the computerized Kiddie-Schedule for Affective Disorders–5 (K-SADS) based on endorsement from either child- or parent-report. A binary diagnosis of whether participants have ever had a K-SADS DEP diagnosis up to year 2 of the ABCD study was used to validate and select the best performing PRS for each ancestry group. Subsequent analysis used DEP severity in the past six months, which was measured using scores from the “Depressive Problems” subscale of the Child Behavior Checklist (CBCL), a widely used measure of emotional problems and provides standardized scores based on national norms in children ages 6–18. The Depressive Problems subscale comprises items consistent with the DSM-5 criteria for DEP and was available from the parent report CBCL and not from the youth. The CBCL has been found to be valid and reliable for children with diverse backgrounds (Albores-Gallo et al., 2007).

### Fitbit Data Quality Control and Summary Statistics

Participants in the ABCD study were invited to participate in an additional 3-week Fitbit study protocol. Daily and weekly summaries of physical activity and sleep were available, which includes total and average metrics by day such as the number of sedentary minutes or average resting heart rate during the daytime. This study utilized the daily Fitbit summary data, which the ABCD team had conducted preliminary quality control (QC) by pre-filtering daily sleep data to days with at least 300 minutes in nighttime wear time. Additional QC measures were performed by this study on the activity data following recommendations of ABCD Fitbit pilot study results (Wing et al., 2022). Days of activity data with less than 600 minutes in daytime wear time were excluded. Participants with less than 7 days of data or with data during Covid-19 starting on March 15th, 2020, were removed. Data was also restricted to the 3-week protocol period by removing additional days of data over 21 days, such that participants included in the analyses had a range of days with data between 7 days and 21 days (Table 1).

**Table 1.** Demographic information for ABCD participants in the whole dataset and in each ancestry that passed quality control for Fitbit data and have valid PRS.

| Demographics | Whole Dataset<br>(n = 2,768) | AFR<br>(n = 350) | AMR<br>(n = 405) | EUR<br>(n = 2,013) |
| --- | --- | --- | --- | --- |
| CBCL DEP Severity Score, Mean (SD) | 53.67 (5.87) | 53.3 (5.89) | 53.8 (5.96) | 53.7 (5.85) |
| Gender, n (%) |  |  |  |  |
| Cis female | 1,366 (49%) | 189 (54%) | 190 (47%) | 980 (49%) |
| Cis male | 1,402 (51%) | 160 (45%) | 215 (53%) | 1027 (50%) |
| Gender Diverse | 7 (1%) | 1 (1%) | 0 (0%) | 6 (1%) |
| Age (in Months), Mean (SD) | 143.42 (7.85) | 143.0 (7.51) | 142.0 (7.97) | 144.0 (7.86) |
| Income-to-Needs Ratio, Mean (SD) | 3.21 (1.85) | 1.9 (1.63) | 1.91 (1.57) | 3.71 (1.69) |
| # of Days with Fitbit Activity Data, Mean (SD) | 21.04 (2.21) | 20.7 (2.39) | 20.9 (2.27) | 21.1 (2.16) |
| # of Days with Fitbit Sleep Data, Mean (SD) | 14.49 (4.24) | 12.8 (4.07) | 14.2 (4.3) | 14.8 (4.19) |
| Mean Daily Daytime Minutes with Fitbit Data, Mean (SD) | 864.3 (48.64) | 884.0 (52.0) | 866.0 (46.8) | 861.0 (47.6) |
| Mean Total Steps, Mean (SD) | 9578.78<br>(3055.73) | 9315.0<br>(2973.0) | 8899.0<br>(3023.0) | 9762.0<br>(3055.0) |
| Mean Total Sleep, Mean (SD) | 449.81 (36.28) | 423.0 (41.2) | 447.0 (35.9) | 455.0 (33.2) |

Summary statistics variables were constructed from daily-level time-series Fitbit data for the main analyses. Participant data were summarized using the average and variance statistics.

Each variable was z-score standardized to facilitate comparison of effect sizes and coefficients across variables, then utilized for mixed effects model analyses. To account for day-to-day variations in total wear time or total sleep time, variables’ values proportional to total wear time (proportion) were created for main analyses, such as the percentage of total sleep time spent in deep sleep (mean proportion deep sleep minutes) or the percentage of total daytime wear minutes spent in sedentary time (mean proportion sedentary minutes).

### GWAS Summary Statistics & Polygenic Risk Scores

GWAS summary statistics for DEP were obtained from meta-analyses and separate outputs for EUR, AFR, and AMR individuals (Wray 2018, Meng 2024). QC was conducted using the GenoPred pipeline on the GWAS summary statistics by only using high-quality variants with a minor allele frequency > 0.01, without ambiguity, no out-of-bound p-values, and no duplicates (Pain et al., 2024).

QC-ed GWAS summary statistics from EUR, AFR, and AMR were used as inputs into the PRS-CS and PRS-CSx PRS methods. PRS-CS is a method that refines single nucleotide variant (SNP) effect sizes using continuous shrinkage on a single set of GWAS summary statistics while accounting for Linkage Disequilibrium (LD) using LD reference panels from the 1000 Genomes Project (Ge et al., 2019). The results of the refined posterior SNP effect sizes were then inputted into PLINK’s linear scoring command to obtain PRS as Ge et al. (2019) suggested and z-scored standardized within each ancestry. The PRS outputs of PRS-CS will hereby be referred to as PRS-CS EUR, PRS-CS AFR, and PRS-CS AMR. PRS-CSx is the successor of PRS-CS by utilizing multiple ancestry-specific GWAS summary statistics to improve cross-population polygenic prediction (Ruan et al., 2022). PRS-CSx provides both ancestry-specific SNP effect sizes outputs (PRS-CSx EUR, PRS-CSx AFR, and PRS-CSx AMR) and a meta option (PRS-CSx META) where an inverse weighted variance analysis is conducted on PRS-CSx EUR, PRS-CSx AFR, and PRS-CSx AMR outputs without fine-tuning on a target population. In total, 7 outputs from PRS-CS and PRS-CSx were compared in a validation dataset that includes ABCD participants without valid Fitbit data to select a single best-performing PRS for each ancestry before subsequent analysis with Fitbit variables and CBCL DEP severity.

### Statistical Analyses

Mixed effects models, as implemented in the GENESIS package in R, were used to investigate associations among DEP-PRS, Fitbit variables, and CBCL DEP severity (Gogarten et al., 2019). Covariates included age, gender, income-to-needs ratio, ancestry principal components (PC), and a genetic relatedness matrix. Genetically derived ancestry PCs and genetic relatedness matrix were derived from a previous study that used the GENESIS package’s pipeline for Principal Components Analysis (PCA) and kinship coefficients accounting for population structures in ABCD subjects (Hyat et al., 2025). Income-to-needs ratio was derived using the income bracket and family size information on each ABCD participant by dividing the mean of the selected income bracket by the federal poverty threshold depending on family size. Our primary analysis using mixed effects models involved three specific setups. First, DEP-PRS derived by PRS-CS and PRS-CSx were tested against lifetime-DEP in ABCD participants (n=7,681) to validate and pick the best-performing PRS for each ancestry. Then, the optimized DEP-PRS were tested for main effects with each Fitbit variable as the dependent variable, including or excluding participants above the subclinical threshold for CBCL DEP severity. Third, the main and interaction effects of each Fitbit variable and DEP-PRS were tested in the same model with CBCL DEP severity as the dependent variable. To account for multiple testing effects, false discovery rate (FDR) was applied to models separately by the type of main analyses and by either activity or sleep. For variables with significant interaction effects, follow-up analyses were conducted within each stratified genetic risk group, defined by low risk = z-scored DEP-PRS < -1, medium risk = z-scored DEP-PRS <= 1, >= -1, and high risk = z-scored DEP-PRS > 1, to explain interaction results rather than using formal pairwise comparisons since there is no established risk categories based on PRS. Ancestry-specific analyses were conducted for the second and third part of the primary analyses. Sensitivity analyses were performed by excluding socioeconomic status as a covariate for ancestry-specific models due to significant differences in income-to-needs ratio between ancestries.

## Results

### Demographics

2,768 participants with valid Fitbit activity and sleep data, CBCL DEP severity assessment at year 2, genotype data, and EUR (n=2,013), AFR (n=350), and AMR (n=405) ancestry were included (Table 1). Participants who passed Fitbit quality control protocol had an average 21.04 days of activity data (SD=2.21) and 14.49 days of sleep data (SD=4.24).

### DEP-PRS, lifetime history of DEP, and current DEP severity

DEP-PRS outputs from PRS-CS and PRS-CSx were compared by testing for association with KSADS lifetime DEP (Table 2). For EUR, PRS-CS EUR (OR = 1.32, p < 0.001, 95% CI [1.19, 1.46]) outperformed PRS-CSx outputs. For AFR, PRS-CSx EUR (OR = 1.14, p = 0.13, 95% CI [0.96, 1.35]) outperformed other PRS-CSx and PRS-CS outputs. For AMR, PRS-CSx META (OR = 1.23, p = 0.02, 95% CI [1.01, 1.50]) outperformed other PRS-CSx and PRS-CS outputs.

**Table 2.**
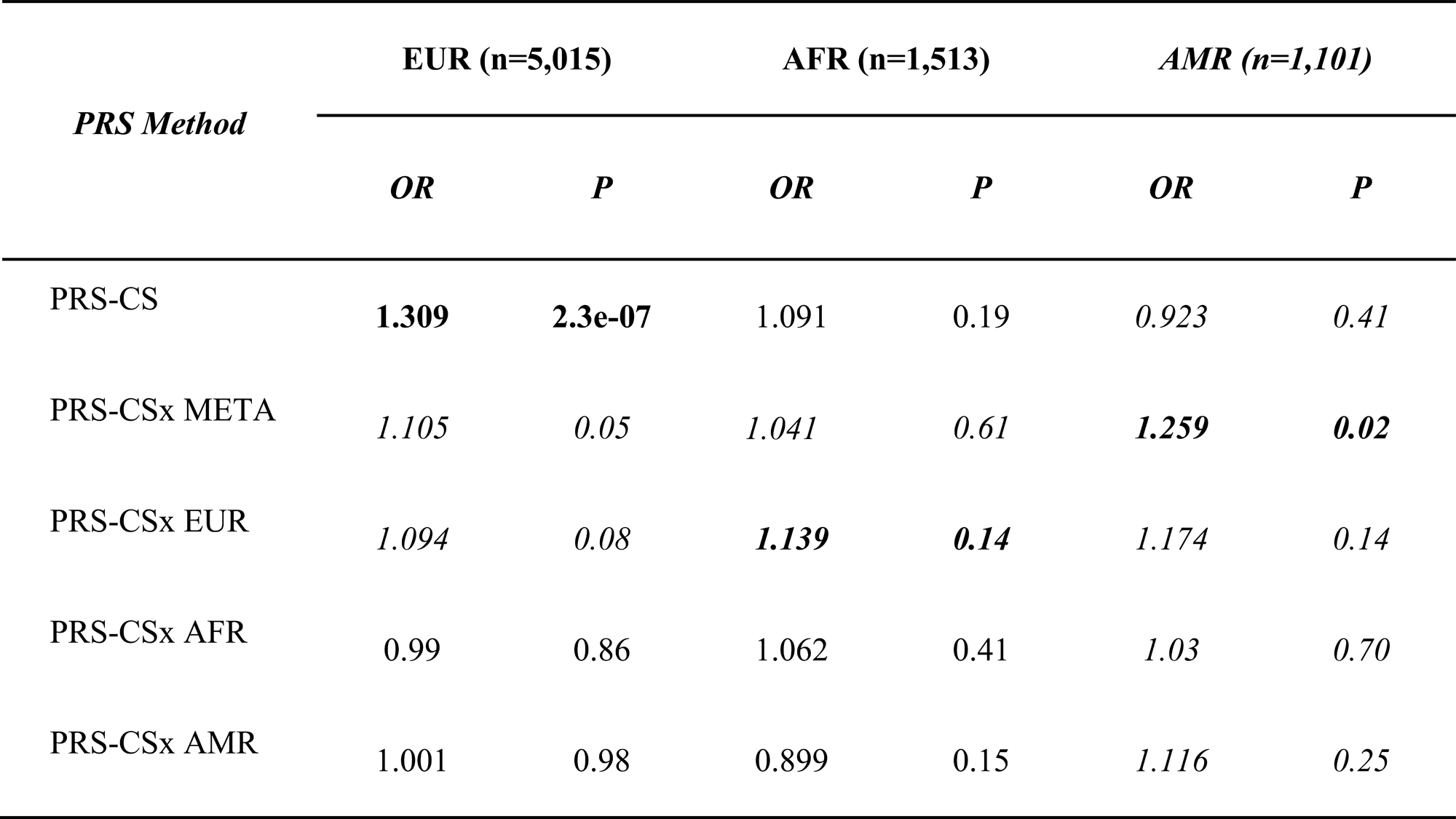
Single-ancestry PRS method (PRS-CS) utilized with each ancestry’s specific GWAS summary statistics was compared against multi-ancestry PRS method (PRS-CSx) that used all 3 GWAS summary statistics in a large ABCD validation sample that includes participants without valid Fitbit data. OR represents the change in odds of KSADS lifetime history of DEP diagnosis per one standard deviation increase in the PRS. KSADS Lifetime History of DEP diagnoses is defined by having at least 1 diagnosis of DEP from baseline up to year 2 of ABCD study. P refers to nominal p value. The best-performing PRS for each ancestry (in bold) were selected to use in association with Fitbit wearable measures and CBCL DEP severity in the main statistical analyses.

### Associations between DEP-PRS and activity and sleep metrics

With each activity measure as the dependent variable, significant associations were observed between DEP-PRS and 9 out of 14 activity measures, 8 of which survived FDR correction. Higher DEP-PRS was associated with higher mean resting heart rate and mean proportion of sedentary minutes, and lower mean total steps and mean and variance of fairly and very active minutes (Figure 1.1A).

**Figure 1.1.**
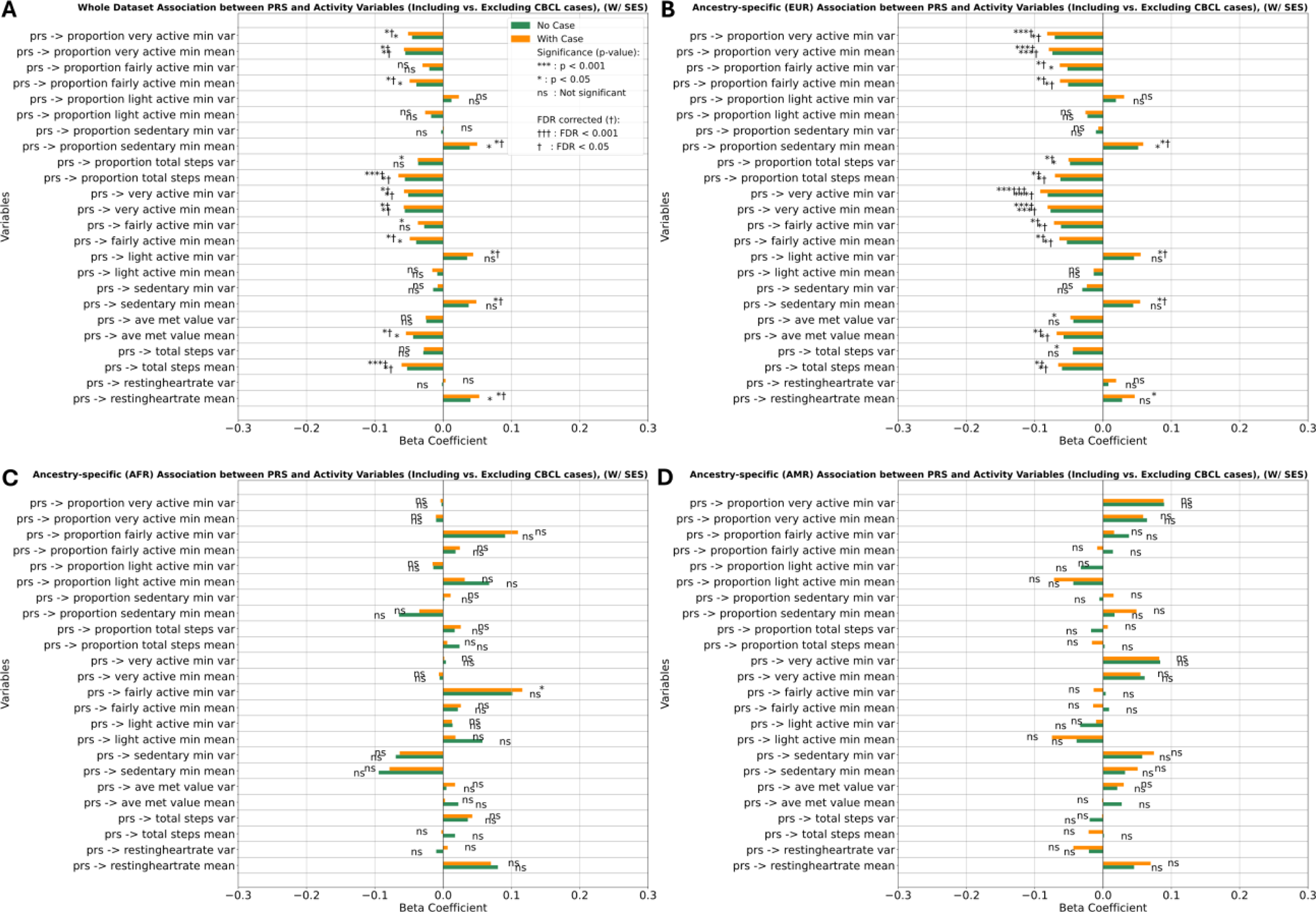
Results of association between DEP-PRS and each Fitbit activity variable as the dependent variable, with FDR correction applied across all models at once. Each association was conducted with all participants included as well as excluding participants who had CBCL DEP Severity score above the designated clinical cut-off score (>= 65), where the participant would be considered as a DEP case. W/ SES refers to the model including income-to-needs ratio as a covariate alongside with age, gender, ancestry PC, and GRM for random effects. The 4 plots include A (Whole dataset), B (EUR), C (AFR), and D (AMR).

With each sleep measure as the dependent variable, significant associations were observed between DEP-PRS and 15 out of 20 sleep measures, 6 of which survived FDR correction (Figure 1.2A). Higher DEP-PRS was associated with lower mean proportion of wake counts and multiple sleep minutes mean measures including total, wake, light, and deep sleep. Higher DEP-PRS was also associated with greater variance in multiple sleep minutes measures including variance in total, light, and REM sleep. DEP-PRS was not associated with any sleep mean or variance measures of proportion in specific sleep stages.

**Figure 1.2.**
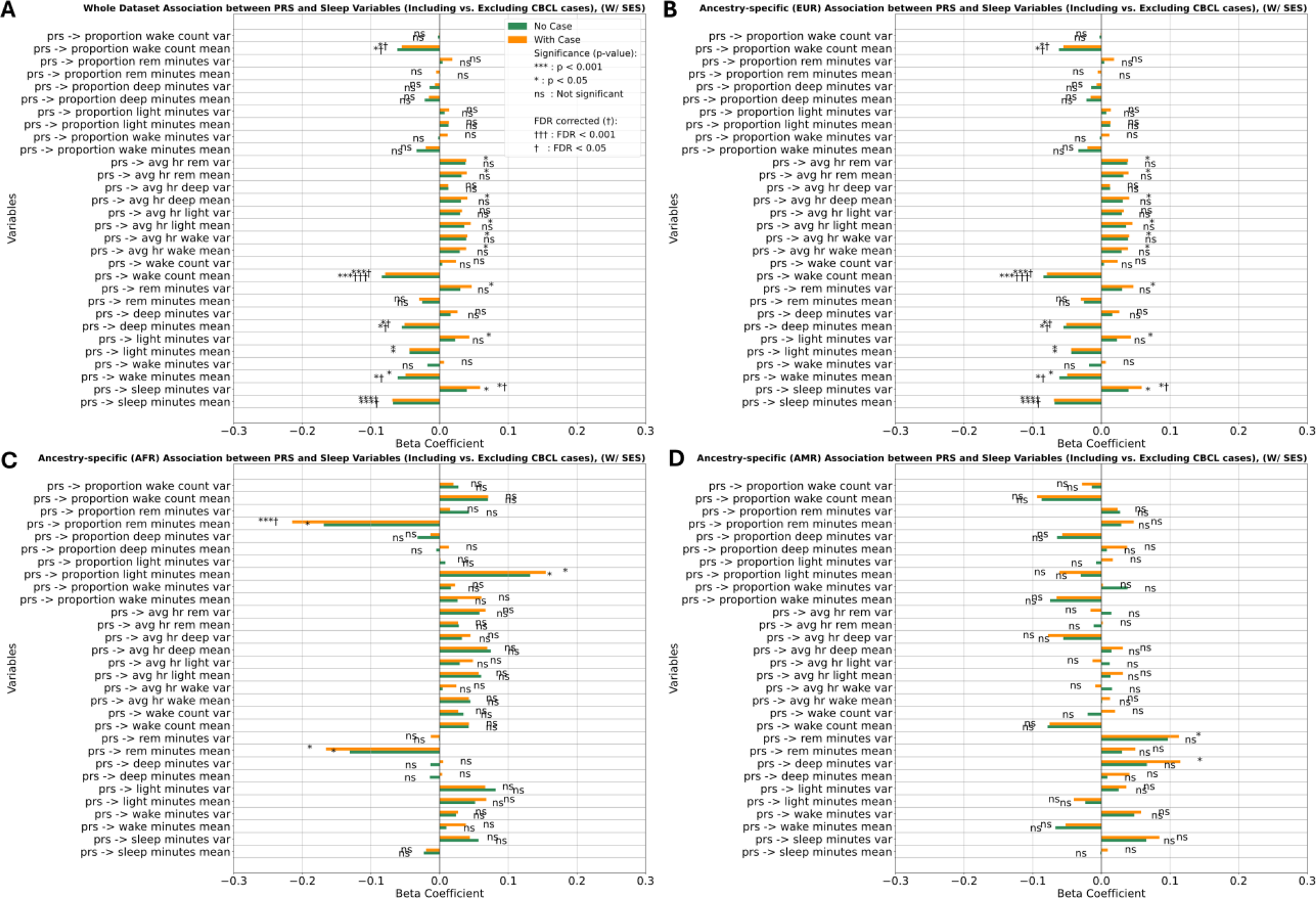
Results of association between DEP-PRS and each Fitbit sleep variable as the dependent variable, with FDR correction applied across all models at once. Each association was conducted with all participants included as well as excluding participants who had CBCL DEP Severity score above the designated clinical cut-off score (>= 65), where the participant would be considered as a DEP case. W/ SES refers to the model including income-to-needs ratio as a covariate alongside with age, gender, ancestry PC, and GRM for random effects. The 4 plots include A (Whole dataset), B (EUR), C (AFR), and D (AMR).

### Associations between DEP severity, DEP-PRS, activity, and sleep metrics

#### Activity

In models for each activity measure and DEP-PRS in the same model where CBCL DEP severity was the dependent variable, DEP-PRS was associated with DEP severity in all models (mean beta (SD) = 0.49 (0.02), FDR p < 0.001) and significant associations were observed between ten out of 14 Fitbit activity variables and CBCL DEP severity, nine of which survived FDR correction (Figure 2.1A). Among the seven significant mean variables, proportion of total steps, sedentary minutes, resting heart rate, proportion of fairly active minutes showed the highest magnitude of effects. For the mean measures of different categories of activity intensity, fairly active minutes had the highest negative association, followed by lightly active minutes and very active minutes. The mean of sedentary minutes, very active minutes, and lightly active minutes each had significant associations while the variance of the respective variables had small effects and non-significance. For variance variables, proportion of fairly active minutes, total steps, and resting heart rate had significant associations.

**Figure 2.1.**
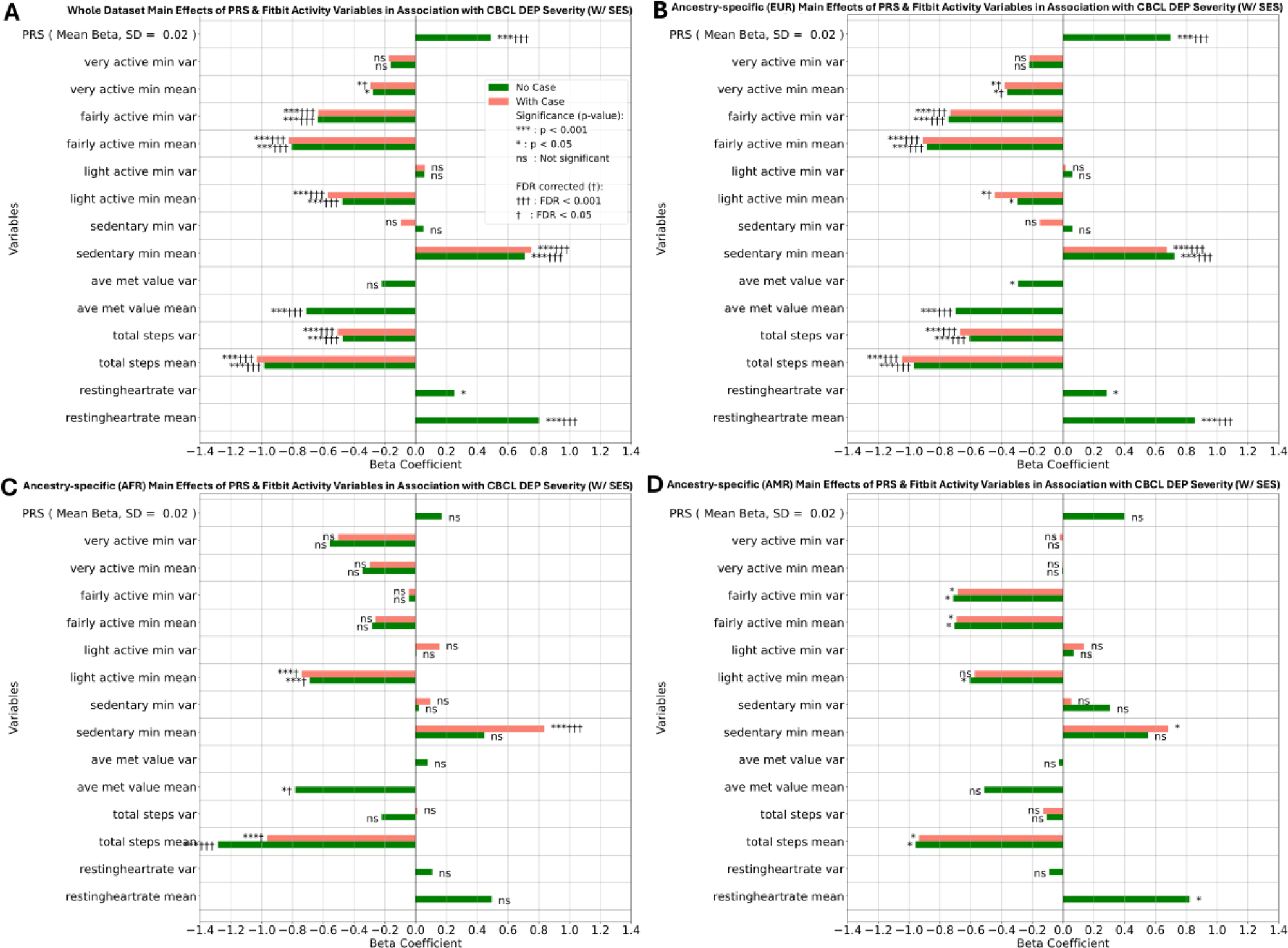
Results of main effects of DEP-PRS and each Fitbit activity variable in association with CBCL DEP Severity. The mean and SD beta for DEP-PRS is calculated with the betas of DEP-PRS across different models, where each model has one Fitbit activity measure and DEP-PRS. Interaction effects were also included in each model, and the results are shown in Figure 2.2. FDR correction was applied across all main and interaction effects of activity measures at once. W/ SES refers to the model including income-to-needs ratio as a covariate alongside with age, gender, ancestry PC, and GRM for random effects. The 4 plots include A (Whole dataset), B (EUR), C (AFR), and D (AMR).

Significant interactions were also observed between multiple activity measures and DEP-PRS, including fairly and very active minutes variance, with the former surviving FDR correction (Figure 2.2A). Follow-up analyses within PRS risk groups for very active minutes variance showed a positive association with DEP severity for subjects in the low risk group and negative associations for subjects in the medium or high PRS group (Figure 2.3A).

**Figure 2.2.**
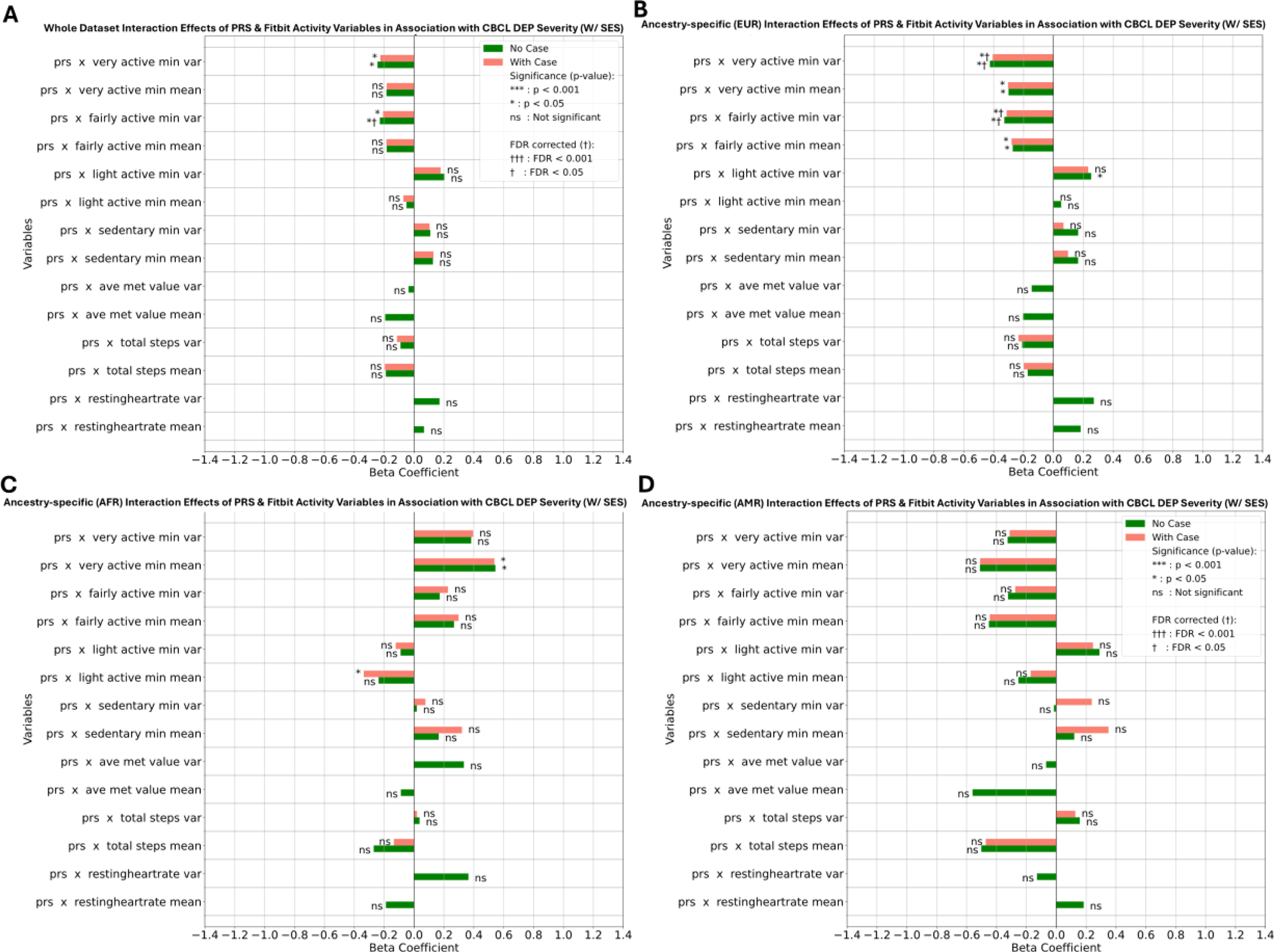
Results of interaction effects of DEP-PRS and each Fitbit activity variable in association with CBCL DEP Severity. Follow-up analyses for interaction effects by PRS risk group are in Figure 2.3. W/ SES refers to the model including income-to-needs ratio as a covariate alongside with age, gender, ancestry PC, and GRM for random effects. The 4 plots include A (Whole dataset), B (EUR), C (AFR), and D (AMR).

**Figure 2.3.**
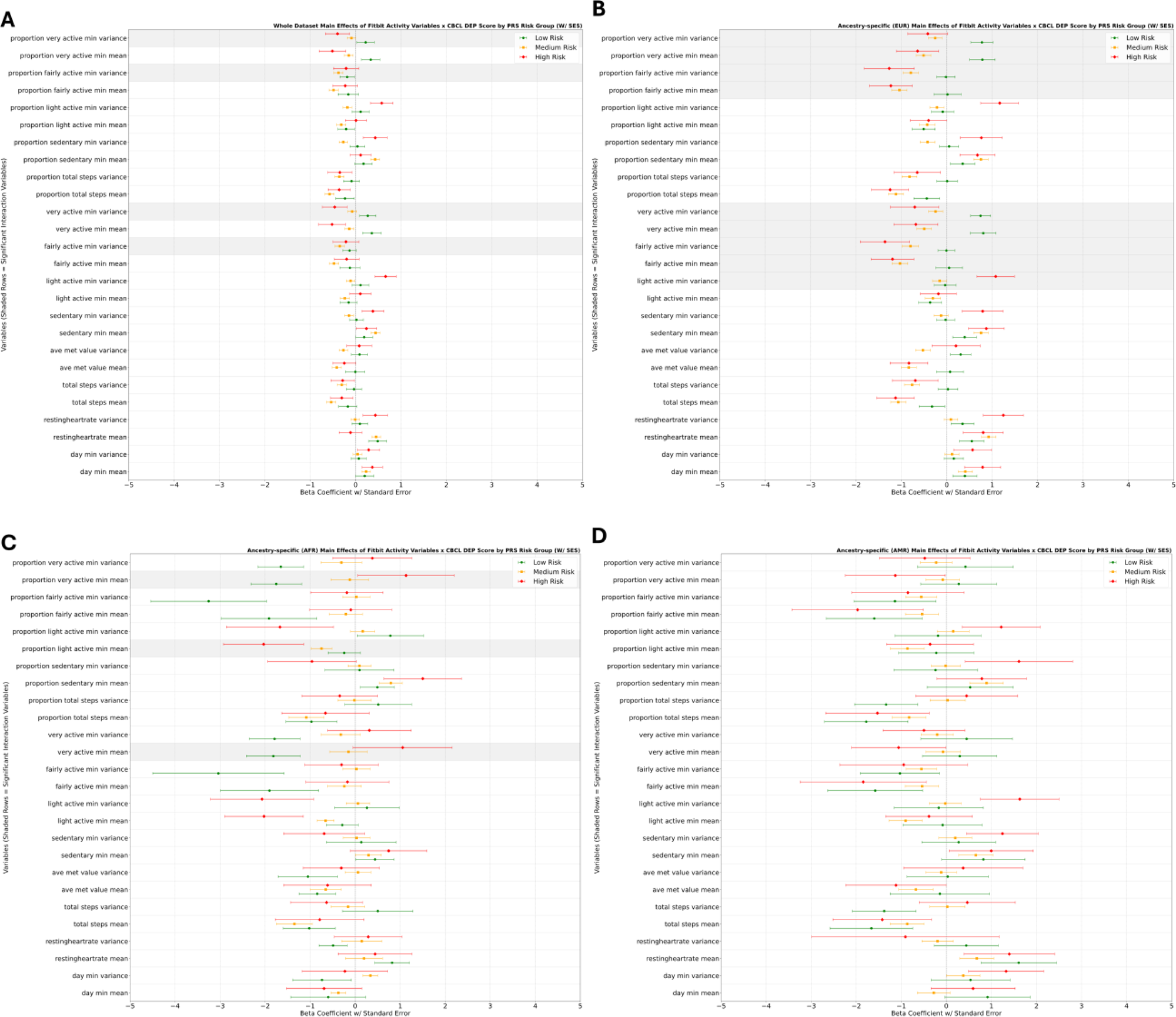
Follow-up analyses of interaction effects with results of main effects between each Fitbit activity measure and CBCL DEP severity within each PRS risk group (low, medium, high). NOTE: no formal pairwise comparisons were conducted, as these analyses are meant for checking interaction results. Activity variables that had significant interactions with DEP-PRS were indicated by shaded rows. W/ SES refers to the model including income-to-needs ratio as a covariate alongside with age, gender, ancestry PC, and GRM for random effects. The 4 plots include A (Whole dataset), B (EUR), C (AFR), and D (AMR).

#### Sleep

In models with each sleep measure and DEP-PRS in the same model where CBCL DEP severity was the dependent variable, DEP-PRS and DEP severity were also significant associated, (mean beta (SD) = 0.49 (0.01), FDR p < 0.001), and significant associations were observed for 15 out of 20 Fitbit sleep variables, 13 of which survived FDR correction (Figure 3.1A). The highest magnitudes of effects were observed in mean heart rate during various sleep stages, variance in light, wake, or total sleep minutes, and variance and mean in wake counts. For the sleep variables in different sleep stages, associations were observed in all of the respective variance variables, while the mean measures for time spent in wake or total sleep were significant. The variance in time spent in light, REM, and deep sleep stages all had higher main effect sizes compared to their respective proportional variables.

**Figure 3.1.**
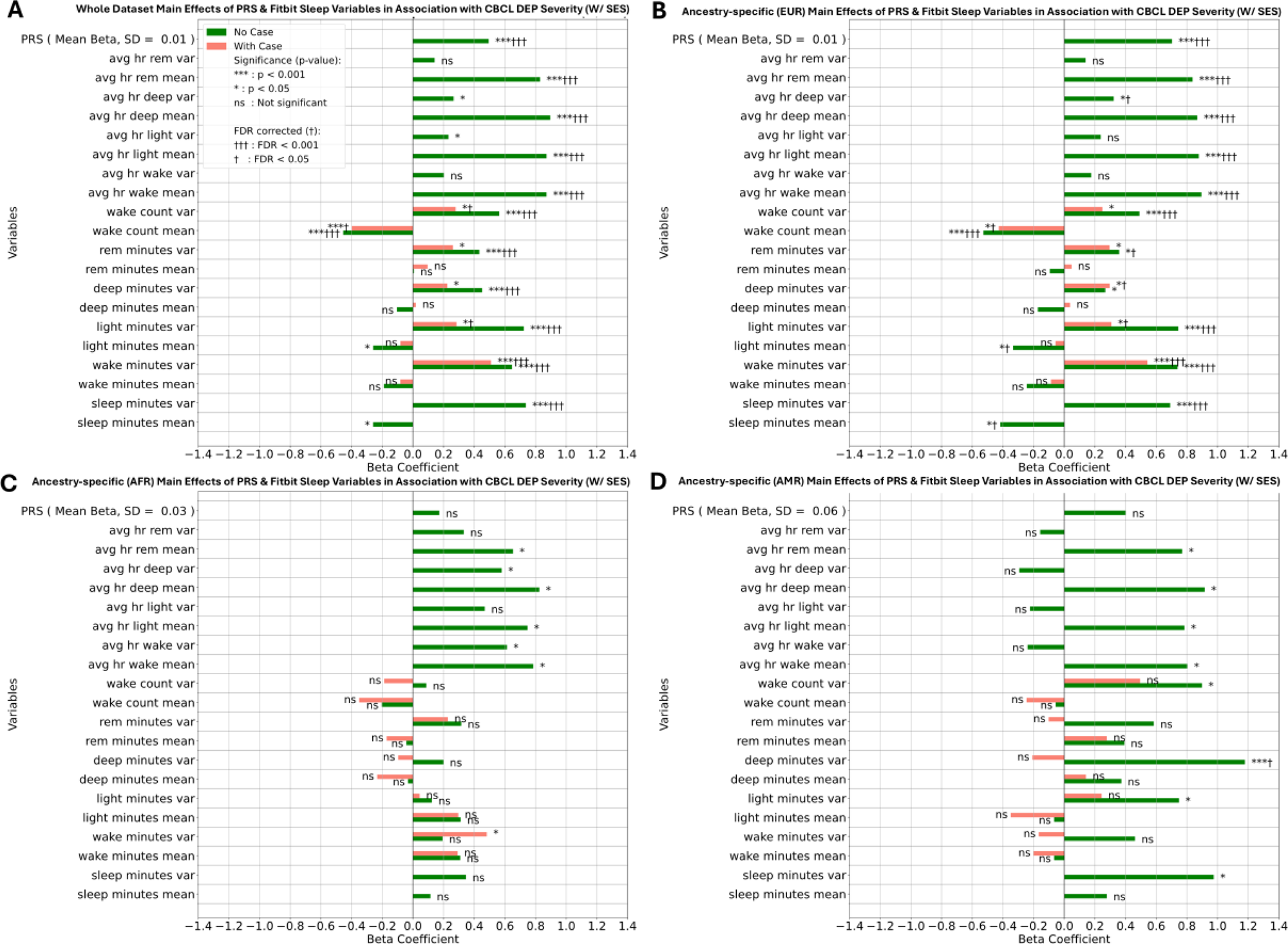
Results of main effects of DEP-PRS and each Fitbit sleep variable in association with CBCL DEP Severity. The mean and SD beta for DEP-PRS is calculated with the betas of DEP-PRS across different models, where each model has one Fitbit sleep measure and DEP-PRS. Interaction effects were also included in each model, and the results are shown in Figure 3.2. FDR correction was applied across all main and interaction effects of sleep measures at once. W/ SES refers to the model including income-to-needs ratio as a covariate alongside with age, gender, ancestry PC, and GRM for random effects. The 4 plots include A (Whole dataset), B (EUR), C (AFR), and D (AMR).

In addition, four interactions between DEP-PRS and sleep metrics for current DEP severity were significant (Figure 3.2A). In particular, there were interactions between DEP-PRS and variances in time spent in each of the four sleep stages, including proportional measures. Follow-up analyses within PRS risk groups showed the positive association between each sleep measure and CBCL DEP severity were stronger for subjects in the high PRS group, followed by the medium and low PRS group (Figure 3.3A). In the high PRS group and among the different sleep stages, variances in proportion of deep sleep minutes had the highest association with DEP severity, followed by wake, light sleep, and rem sleep minutes.

**Figure 3.2.**
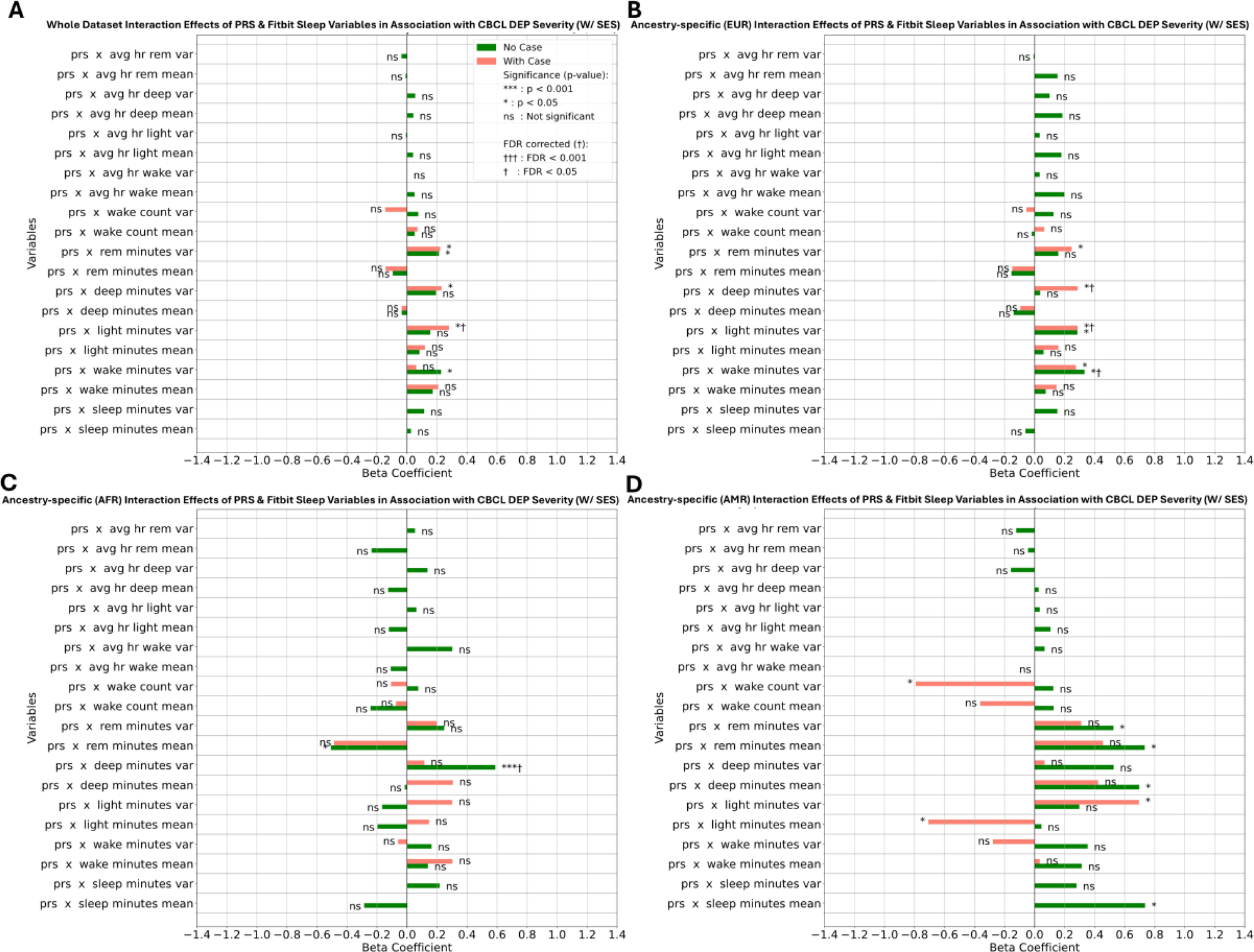
Results of interaction effects of DEP-PRS and each Fitbit sleep variable in association with CBCL DEP Severity. Follow-up analyses for interaction effects by PRS risk group are in Figure 3.3. W/ SES refers to the model including income-to-needs ratio as a covariate alongside with age, gender, ancestry PC, and GRM for random effects. The 4 plots include A (Whole dataset), B (EUR), C (AFR), and D (AMR).

**Figure 3.3.**
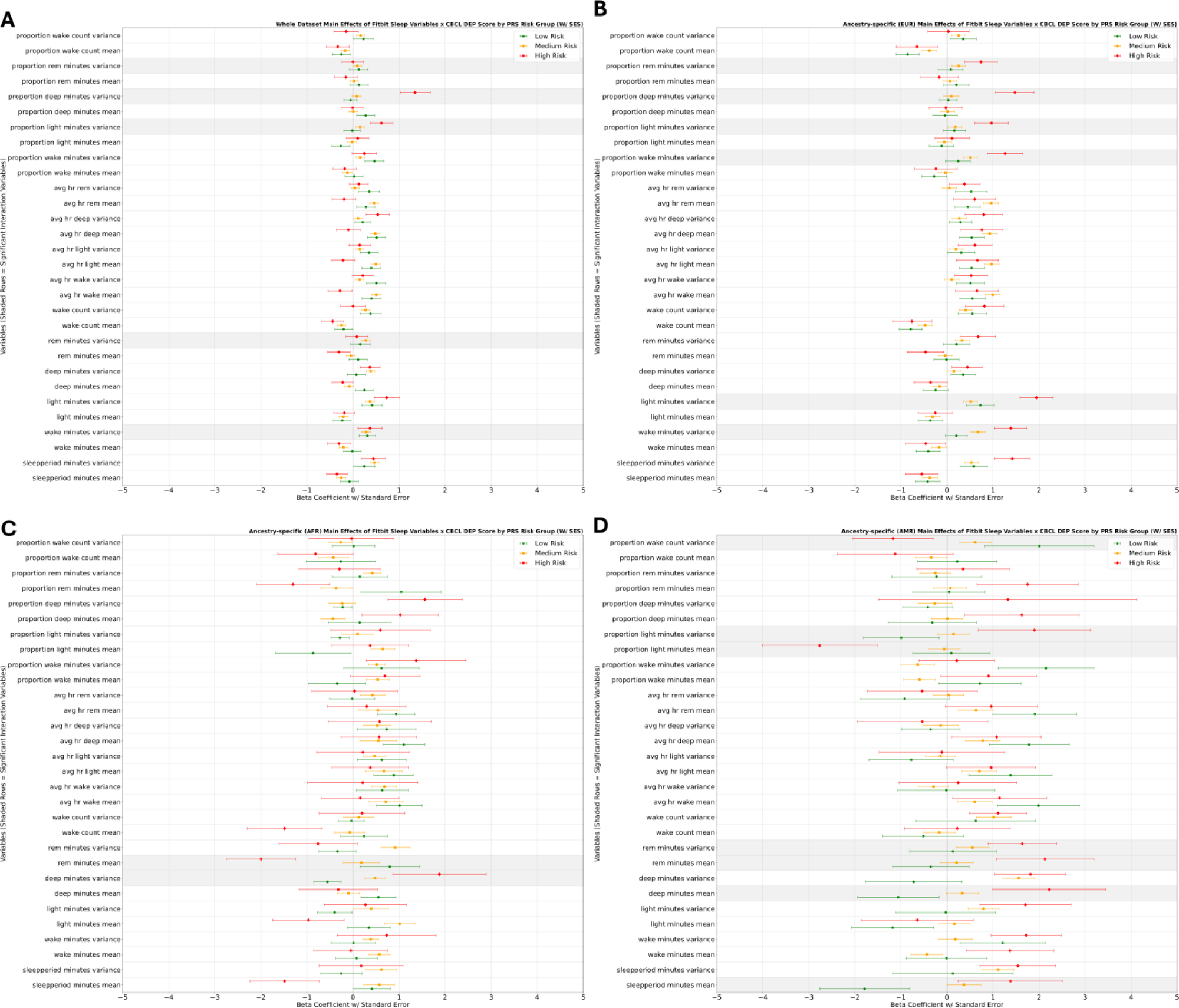
Follow-up analyses of interaction effects with results of main effects between each Fitbit sleep measure and CBCL DEP severity within each PRS risk group (low, medium, high). NOTE: no formal pairwise comparisons were conducted, as these analyses are meant for checking interaction results. Sleep variables that had significant interactions with DEP-PRS were indicated by shaded rows. W/ SES refers to the model including income-to-needs ratio as a covariate alongside with age, gender, ancestry PC, and GRM for random effects. The 4 plots include A (Whole dataset), B (EUR), C (AFR), and D (AMR).

### Ancestry-stratified associations between DEP severity and DEP-PRS, activity, and sleep metrics

#### Activity

Significant associations detected between DEP-PRS and activity measures in the multi-ancestry model became stronger and had higher effect sizes in the EUR only sample (Figure 1.1B). For AFR, the only significant association was between PRS and increased fairly active minutes variance, which was in the positive direction compared to the significant negative association for EUR (Figure 1.1C). For AMR, no significant associations were found (Figure 1.1D).

Significant main associations between each Fitbit activity variable and CBCL DEP severity remained significant with similar effect sizes in the EUR only sample (Figure 2.1B). In both AFR and AMR, there were less significant associations, but effect directions remained similar (Figure 2.1C & Figure 2.1D).

Significant interactions from the multi-ancestry model that remained significant in the EUR only sample included light, fairly, and very active minute variance, while the interactions for the mean of the respective variables became significant (Figure 2.2B). For AFR, nominally significant interactions include PRS and very active minutes mean, which was in the opposite, positive direction compared to EUR, as well as a negative interaction with light active minutes mean, which were not found in EUR or AMR (Figure 2.2C & Figure 2.2D). Follow-up analyses suggested the association between very active minutes mean and DEP severity for AFR is positive for the high risk group and negative for the low risk group, and vice versa for the EUR follow-up analyses (Figure 2.3B & Figure 2.3C). Furthermore, follow-up analyses suggested the association between light active minutes mean and DEP severity for AFR was negative only for the high risk group. For AMR, no significant interactions were observed, while effect sizes were generally more similar to EUR rather than AFR (Figure 2.3D).

#### Sleep

Significant associations between DEP-PRS and sleep measures in the multi-ancestry sample became less significant but had similar directions and effect sizes in the EUR only sample (Figure 1.2B). For AFR, 2 significant associations were observed between DEP-PRS and decreased mean proportion of REM sleep minutes, which survived FDR correction, and increased mean proportion of light sleep minutes (Figure 1.2C). Both significant associations in AFR were not observed in full sample, EUR, or AMR, and had higher effect sizes compared to significant variables in other ancestries. For AMR, DEP-PRS was nominally positively associated with deep and REM sleep variance, where the latter association was observed in the full and EUR only samples (Figure 1.2D).

Significant main associations between each sleep variable and CBCL DEP severity remained significant with similar effect sizes in the EUR only sample (Figure 3.1B). The significant negative associations of total and light sleep minutes mean with DEP severity were only observed in EUR. The effect sizes for those variables were in the opposite direction but non-significant for other ancestries, specifically with light minutes mean for AFR and total sleep minutes mean for AMR (Figure 3.1C & Figure 3.1D). Significant associations between sleep variance measures and DEP severity were in similar, positive directions for EUR and AMR, but stronger effect sizes were observed for AMR with total and deep sleep variance.

Significant interactions from the multi-ancestry model, specifically sleep variance measures, remained significant in the EUR sample (Figure 3.2B). Follow-up analyses for EUR suggest similar associations to the multi-ancestry sample as well (Figure 3.3B). For AFR, a nominally significant negative interaction between PRS and REM sleep minutes mean was observed, which was not observed in EUR and observed as an opposite, positive interaction for AMR (Figure 3.2C & Figure 3.2D). Follow-up analyses suggested that the association between REM minutes mean and DEP severity is strong and negative for the AFR high risk group, and negative for the AMR high risk group (Figure 3.3C & Figure 3.3D). For AMR, 4 nominally significant associations, including positive interactions with total and deep sleep minutes mean and negative interactions with light sleep minutes mean and wake counts variance, were not observed in other ancestries nor in the full sample. Follow-up analyses suggested positive associations between total and deep sleep minutes mean and DEP severity for the AMR high risk group, and negative associations for both variables in the AMR low risk group.

## Discussion

This study examined the interplay among DEP-PRS, wearable-based physical activity and sleep data, and DEP severity outcome measures. DEP-PRS was associated with lifetime DEP, activity, and sleep measures. DEP-PRS also showed interaction effects with wearable measures of activity and sleep in association with DEP severity, where stronger associations between wearable measures and DEP severity were observed in individuals at higher genetic risk. There are ancestry-specific differences in wearable associations in individuals at higher genetic risk.

Recent literature has suggested multi-ancestry methods, like PRS-CSx, could perform better than single-ancestry methods on diverse populations depending on method optimization, GWAS data selection, and ancestral population, but have not utilized psychiatric phenotypes (Wang et al., 2023; Gunn et al., 2025). Several studies on the ABCD dataset have associated multi-ancestry PRS and DEP measures using PRS-CS and the PRS-CSx META option (Ahern et al., 2023, Lee et al., 2025). This study corroborates the stronger association between PRS-CS EUR and KSADS lifetime DEP for EUR in the ABCD dataset but also indicates the ancestry-specific outputs of PRS-CSx, which were not used by previous studies, could be useful. PRS-CSx EUR outperformed both PRS-CSx META and PRS-CS AFR for AFR, while PRS-CSx META performed best for AMR. Alternatively, another study on the ABCD dataset used the same KSADS outcome measure and derived multi-ancestry PRS with SBayesRC, a PRS method that integrates functional annotations to pinpoint causal variants (Xu et al., 2024, Zheng et al., 2024). Xu et al. derived odds ratios for EUR and multi-ancestry populations at 1.33 ([1.15 – 1.52], p<0.001) and 1.31 ([1.15 – 1.50], p<0.001), respectively, while this study found odds ratios for the same outcome at 1.31 ([1.18 - 1.45], p<0.001) and 1.20 ([1.11 - 1.29], p<0.001). However, this study utilized ancestry-specific GWAS summary statistics to take advantage of the fine-mapping approach of PRS-CSx, while Xu et al. (2024) utilized a different set of meta-analyzed GWAS summary statistics (Major Depressive Disorder Working Group of the Psychiatric Genomics Consortium, 2025). Therefore, future studies are needed to systematically evaluate the use of existing PRS methods and GWAS summary statistics for diverse populations with psychiatric phenotypes, as more polygenicity and heterogeneity could implicate PRS accuracy.

In a previous systematic review of wearable features in association with DEP, 22 of 48 studies reported statistically significant associations for physical activity variables, with vigorous activity having one of the higher associations among all reported variables (Rohani et al., 2018). While studies from Rohani et al. (2018) were adult samples, recent studies using the Fitbit measures from the ABCD dataset have indicated higher mean resting heart rate, lower mean and variability in total steps, and lower average time spent in fairly and very active minutes are associated with higher internalizing symptoms (Nelson et al., 2022; Damme et al., 2024). Results from this study corroborate the main effects in association with CBCL DEP severity among all previously mentioned variables and increased mean sedentary minutes. Extending on previous studies, this study identified significant interaction effects in the multi-ancestry sample between PRS and average total steps and variance in fairly and very active minutes, with stronger negative associations as PRS increases. Existing studies on adult populations have found no interactions between a EUR-only PRS and self-report activity and indicate physical activity as a protective factor for DEP risk across different levels of genetic predisposition (Choi et al., 2020, Zhao et al., 2023). However, our study used multi-ancestry optimized PRS and objective measures of behavior and identified cross-sectional significant interactions. A previous study also suggests higher rates of vegetative symptoms in adolescent-onset DEP compared to adult recurrent DEP (Rice et al., 2019). Results of this study further indicate a need for longitudinal studies to examine the bidirectional relationship between lifestyle factors and DEP in adolescents depending on genetic risk. Alternatively, in ancestry-specific analysis, the association between mean very active minutes and DEP severity was more positive as PRS increases for AFR, but the opposite interaction effects were identified for EUR. This discordant association suggests caution in interpreting wearable-derived minutes of intense activity, as such measures can capture both potentially protective physical activity and activity changes due to differences in socioeconomic factors (Sallis et al., 1996).

Existing systematic reviews suggest associations between wearable measures of sleep duration variability and sleep efficiency or sleep quality, rather than average sleep duration, with DEP symptoms (Zierer et al.,2024; Messman et al., 2024; De Angel et al., 2022). While the lack of associations for average sleep duration could be due to non-linear associations, a prior study in the ABCD dataset found no significant non-linear association for wearable measures of average sleep duration, and implicates higher variability with higher internalizing symptoms as well (Nelson et al., 2022). Our study is consistent with these prior findings and builds on them, in terms of observing consistent associations of variability in sleep duration, variability in wake time during sleep, as well as mean wake counts with DEP severity, as well as interaction effects w/ PRS, which suggests these variables are generally associated but even more relevant for individuals with higher genetic predisposition. In ancestry-specific analyses, while we found average sleep duration had non-significant main effects, average duration of sleep in in specific sleep stages show significant interactions with PRS, highlighting the importance of average sleep duration in specific sleep stages when considered within-ancestry and alongside PRS due to potential genetic and race/ethnicity factors. Furthermore, our analyses indicate that for the African ancestry population, PRS is negatively associated with the proportion of REM sleep as well as a significant interaction effect between higher REM sleep and higher PRS with higher DEP symptoms, while PRS was not associated with the proportion of REM sleep in the trans-ancestry model or for other ancestries. Meta-analysis of objective sleep measurements in association with DEP, including both sleep efficiency measures and sleep architecture measures, have further emphasized the importance of the sleep variability and efficiency measures, rather than the non-significant effects for total deep sleep, REM sleep, REM density, and REM latency and significant heterogeneity in true effect sizes of these variables across the pooled studies (Lovato & Gradisar, 2014; Augustinavicius et al., 2014). However, these studies were not ancestry-specific, and there are studies on racial and ethnic differences in sleep architecture that warrants further investigation on whether these differences also present as risk factors for DEP (Johnson et al., 2019; Rao et al., 2019, Poland et al., 1999). Future studies should explore whether these sleep differences are risk factors for DEP, utilize ancestry-specific longitudinal analysis, or incorporate more accurate genetic influence measures.

The strengths of this study include the use of a large (n=2,768) and diverse (AFR, AMR, and EUR) population, multi-ancestry PRS methods, and wearable data to derive associations between activity, sleep, and DEP symptoms depending on genetic predisposition and ancestry. However, there are several limitations . First, this is a cross-sectional analysis on the ABCD dataset, where CBCL symptoms were measured right before the Fitbit protocol at year 2, which means causality cannot be established. Second, although multi-ancestry PRS methods performed better than single-ancestry PRS methods, the association between PRS and both KSADS lifetime history of DEP and CBCL DEP severity for the AFR population is still not significant and could hinder the validity of associations with behavior variables, which emphasizes the need for optimized PRS methods. Third, further investigation is needed to compare the accuracy of self-report and wearables with gold-standard measures concurrently, especially when clinical symptoms are present (Kawai et al., 2023). Several studies do indicate significant differences between wearables and gold-standard PSG in measuring sleep when individuals present clinical symptoms of DEP or insomnia (Cook et al., 2017; Kang et al., 2017). The accuracy of wearable measures of macro sleep architecture (light, deep, and REM sleep) is lower than total sleep time and sleep efficiency, which could affect the validity of associations this study found for specific sleep stages (Haghayegh et al., 2019). Also, the accuracy for measuring energy expenditure, which is used to categorize the number of minutes in light, fairly, and very active minutes, is less than total steps (Fuller et al., 2020). Wearables capture all forms of movement and do not automatically infer the specific types of activities. Even though there is a significant negative association between specific intensity of active minutes and DEP severity, active minutes can capture activities that may have different associations with mental health if able to be classified separately (Butte et al., 2018). Furthermore, previous research highlighted that ABCD participants’ socioeconomic demographics were significantly associated with their willingness to participate in the additional Fitbit study and adherence to device wear time, which could induce bias into ABCD wearable prediction studies (Kim et al., 2023).

In conclusion, this study was able to utilize a diverse U.S. adolescent population dataset with genotype data, Fitbit wearable data, and psychiatric assessments to elucidate the gene-environment interactions that warrant further longitudinal studies with research-grade activity and sleep measurements to test the effectiveness of lifestyle interventions for DEP or as predictors of DEP onset.

## Data Availability

All data produced in the present study are available upon reasonable request to the authors.

